# Polyethylene and Polyvinyl Chloride Nanoplastics Accelerate Atherosclerosis Through Distinct Smooth Muscle Cell Phenotypic Transitions

**DOI:** 10.64898/2026.02.10.26345390

**Authors:** Siwen Zheng, Wenduo Gu, Quanyi Zhao, Yoko Kojima, Kaylin C. A. Palm, Michal Mokry, Kai-Uwe Jarr, Hua Gao, Isabella Damiani, Guyu Qin, Gurmenjit Bahia, Sugandha Basu, Ramendra Kundu, Matthew Worssam, João P. Monteiro, Alexa Berezowitz, Chad S. Weldy, Paul Cheng, Gerard Pasterkamp, Nicholas J. Leeper, Juyong Brian Kim

## Abstract

Micro- and nanoplastics (MNPs) are increasingly detected in human tissues, yet their causal contribution to cardiovascular disease remains poorly understood. Here we show that oral exposure to polyethylene (PE) and polyvinyl chloride (PVC) -- the most abundant polymers found in human atheromas -- accelerates atherosclerosis in ApoE-/- mice through distinct, polymer-specific molecular mechanisms. While both polymers increased plaque burden and reduced contractile smooth muscle cell (SMC) markers, single-cell transcriptomic profiling revealed divergent phenotypic trajectories. PE exposure drives SMCs toward a chondromyocyte-like cell (CMC) state, characterized by upregulated osteogenic signaling and markedly increased vascular calcification. Conversely, PVC exposure promotes a fibromyocyte-like program associated with altered collagen metabolism and accelerated cell migration without enhancing calcification. These distinct SMC programs are reflected in the transcriptional signatures of symptomatic human carotid plaques, suggesting clinical relevance for polymer-specific vascular remodeling. Our findings establish a causal link between common environmental plastics and accelerated atherosclerosis, demonstrating that MNP-induced vascular risk is mediated by divergent SMC fate decisions. These results provide a mechanistic framework for assessing the cardiovascular impact of global plastic pollution and identifying potential therapeutic targets to mitigate MNP-associated vascular toxicity.

---

Plastic products are ubiquitous in modern life, and their degradation into micro- and nanoplastics (MNPs) is increasingly recognized as an emerging cardiovascular health concern.^1^ Recent human studies have identified substantial accumulation of polyethylene (PE) and polyvinyl chloride (PVC) MNPs in atherosclerotic plaques and link them to adverse cardiovascular outcomes.^2^ Consistent with prior evidence linking plastic exposure to atherosclerosis, studies in *Apoe-/-*mice showed that oral exposure to polystyrene exacerbates atherosclerotic lesion development.^3^ However, whether the most abundant polymers detected in human plaques (i.e., PE and PVC) lead to atherosclerosis, and the molecular mechanisms by which they perturb vascular homeostasis, remain poorly defined.

To address this, 8-week-old atherosclerosis-prone male *Apoe*?*-/-*? mice were fed a high-fat diet and given either regular drinking water or water containing 100-nm PE or PVC beads (1 μg/mL; n=15 per group) for 16 weeks. This design models oral intake as a major route of human exposure to MNPs. The dose employed here is approximately 10-fold higher than concentrations typically reported in drinking water but remains within the range measured in heavily contaminated regions.^4^ This level was selected to facilitate mechanistic analysis of vascular effects within the limited duration and single-route exposure design of the in vivo study. At the end of the treatment period, PE- and PVC-exposed mice exhibited higher body weight (Figure [Aa]) and systolic blood pressure (Figure [Ab]) relative to controls. Despite unchanged plasma LDL and triglyceride levels in both exposure groups (Figure [Ac-Ad]), the areas of Oil Red O (ORO) positive lipid-rich plaque were increased in both *en face* aortas and the aortic sinus (Figure [Ae] and [Af]). Furthermore, Transgelin (TAGLN) positive plaque area was reduced in the plaques from both PE- and PVC-exposed mice (Figure [Ag]).

**Figure.** Polyethylene and polyvinyl chloride nanoplastics accelerate atherosclerosis through distinct smooth muscle cell phenotypes. *Apoe-/-*mice were exposed to polyethylene (PE) or polyvinyl chloride (PVC) nanoplastic beads (100 nm, 1 μg/mL) via drinking water for 16 weeks under a high-fat diet. The drinking water was refreshed every 2–3 days to minimize time-dependent changes in nanoparticle physicochemical properties and to maintain consistent exposure conditions. **A**, Altered systemic physiology and smooth muscle cell remodeling. **Aa**, Body weight. **Ab**, Systolic blood pressure. **Ac**, Plasma LDL levels. **Ad**, Plasma triglyceride levels. **Ae**, Representative images of *en face* oil red O (ORO) staining of aortas (left) with quantitative analysis of ORO-positive area normalized to total aortic area (right). **Af**, Representative ORO staining of the aortic sinus (left) with quantitative analysis of ORO-positive area normalized to plaque area (right). **Ag**, Representative transgelin (TAGLN) staining of the aortic sinus (left) with quantitative analysis of TAGLN-positive area normalized to plaque area (right). Scale bars in **Af** and **Ag** represent 500 μm. **B**, Single cell transcriptomic characterization of mouse aortic root atherosclerotic plaques. **Ba**, Uniform Manifold and Approximation and Projection (UMAP) visualization of cell types in the mouse aortic root based on an integrated dataset from control, PE, and PVC groups (n=42,163 cells). SMC, contractile smooth muscle cell; FMC, fibromyocyte; CMC, chondromyocyte; AdvFibro, adventitial fibroblast; ValveFibro, valve fibroblast. **Bb**, Numbers of differentially expressed genes (DEGs) across major cell types in PE- and PVC-exposed mouse aortic roots relative to control (n=4 mice per group). SMC populations comprise the combined populations of contractile SMCs, FMCs, and CMCs. **Bc**, Overlap of DEGs in SMC populations following PE and PVC exposure. **Bd**, Volcano plot showing DEGs between PE and PVC groups in SMC populations. FC, fold change. The top 15 genes ranked by FC for PE (green dots) and PVC (green dots) are labeled. **Be**, Proportional distribution of SMC populations across experimental groups. **Bf**, Heatmap of DEG counts across SMC populations. DEGs were first identified separately for PE vs control and PVC vs control within each SMC phenotypes and then overlapped to categorize genes as PE-specific (unique to PE vs control), PVC-specific (unique to PVC vs control), or shared (significant in both comparisons). Within each annotated cell type, DEGs between exposure and control groups (**Bb, Bc**, and **Bf**) were identified using a Wilcoxon rank-sum test (adjusted P < 0.1; cells expressing the gene ≥10%. **Bg**, Gene Ontology (GO) enrichment analysis highlighting pathways preferentially upregulated in CMCs (left) and FMCs (right). FDR, false discovery rate. Count, number of genes annotated to a given GO term. C. Validation in human cohort and human coronary artery smooth muscle cells. **Ca**, Supportive human evidence from patients with severe symptoms from the Athero-Express Biobank. Bubble plot showing selected GO pathways enriched among SMC-associated gene sets derived from mouse aortic single-cell RNA-seq data and evaluated in human carotid plaque transcriptomes from patients with more severe symptoms, comparing PE vs Control and PVC vs Control. Primary human coronary artery smooth muscle cells (HCASMCs) were used for (**Cb-Cf**) *in vitro* phenotypic validation after 24 h exposure to 100 nm PE (50 mg/L) and PVC (10 mg/L) nanoplastic beads, with concentrations selected based on preliminary dose-response experiments identifying the minimal concentrations that induced transcriptional changes. **Cb**, Wound-healing scratch assay, with wound area normalized to the corresponding value at time 0 for each condition. **Cc**, EdU incorporation assay to quantify proliferation, with EdU-positive signal normalized to Hoechst-stained nuclei. **Cd**, Relative expression of *HAPLN1* (Hyaluronan and proteoglycan link protein 1) following 14 days of osteo/chondrogenic induction, normalized to basal medium for each condition. **Ce**, Quantification of calcium content. **Cf**, Representative Alizarin Red S (ARS) staining images (left) with quantitative analysis of ARS-positive area normalized to total area (right). Scale bar represents 640 μm. Statistical significance between groups was assessed by ordinary one-way ANOVA followed by Tukey’s multiple comparisons test. *P < 0.05, **P < 0.01, ***P < 0.001, ****P < 0.001, ns not significant.

To explore whether the observed changes in the plaque characteristics were linked to alterations in specific cell types, we next performed single-cell RNA-sequencing (scRNA-seq) of the aortic root (Figure [Ba]). Analysis of the scRNA-seq revealed SMC populations to have the most pronounced transcriptional response to PE and PVC exposure, as reflected by pseudo-bulk differential expression analyses (Figure [Bb]). Within SMC populations, we observed 450 PE-specific, 846 PVC-specific, and 499 shared differentially expressed genes (DEGs) relative to controls (Figure [Bc]). Differential gene expression testing between PE and PVC groups revealed treatment specific gene expression effects, highlighting *Col2a1, Ibsp, Omd, Cytl1, Pdk4* (upregulated in PE) as well as *Sfrp2, Kif26b, Galnt16, Osbpl3, Gdpd5* (upregulated in PVC) (Figure [Bd]).

During atherosclerosis progression, SMCs undergo transitions from contractile SMCs to fibromyocytes (FMCs) and ultimately to chondromyocyte-like cells (CMCs).^5^ We found that PE and PVC altered SMC differentiation pattern in different ways. Across all SMC phenotypic states, PE increased the CMC fraction by 81.4% (2.7% to 4.9%), whereas no comparable increase was observed following PVC exposure (Figure [Be]). When DEGs were stratified by cell state, PE-specific DEGs were concentrated in CMCs (97 PE-specific vs 20 PVC-specific; 5 shared), while PVC induced a larger state-specific response in contractile SMCs and FMCs (Figure [Bf]). Gene Ontology analysis linked PE-associated changes to chondrogenic/osteogenic pathways, whereas PVC-associated changes were enriched for fibroblast growth factor response, connective tissue development, and cell–substrate adhesion (Figure [Bg]). Notably, the PE-induced (chondrogenic/osteoclast) and PVC-induced (FGF-response/collagen metabolism) SMC programs were recapitulated in human carotid plaques from the Athero-Express (AE) Biobank (>3,500 endarterectomy specimens; local IRB approval), where higher module expression tracked with greater symptom severity, supporting clinical relevance (Figure [Ca]).

We further validated these findings *in vitro* using primary human coronary artery smooth muscle cells (HCASMCs). In wound-healing scratch assays, PE slowed wound closure, whereas PVC accelerated closure (Figure [Cb]). Because scratch closure can reflect both migration and proliferation, we measured proliferation by EdU incorporation. PVC significantly reduced EdU incorporation, while PE had no effect (Figure [Cc]), indicating that the accelerated closure observed with PVC is unlikely to be driven by increased proliferation and is more consistent with altered cell migration. After 14 days of osteo/chondrogenic induction, PE markedly promoted calcification, supported by a significant upregulation of the calcification-associated marker *HAPLN1* (Hyaluronan and proteoglycan link protein 1; Figure [Cd]), along with increased calcium deposition, as quantified by both total calcium content (Figure [Ce]) and Alizarin Red S positive area (Figure [Cf]). In contrast, PVC induced no such increase in calcification across these readouts.

Together, our data show that PE and PVC accelerate atherosclerosis via distinct SMC programs. PE promotes chondrogenic transition and calcification, whereas PVC favors fibroblast-like remodeling and accelerated wound closure despite reduced proliferation. These findings should be interpreted with several limitations. We tested 100-nm spherical beads, while environmental MNPs are often irregular fragments and fibers, which may exhibit different vascular effects. In addition, only male mice were studied, and potential sex-specific effects remain to be determined. Nonetheless, our findings provide a mechanistic framework linking plaque⍰relevant polymers to divergent SMC fate decisions and functional remodeling, thereby motivating future efforts to identify actionable pathways and therapeutic targets to mitigate MNP⍰associated vascular risk.

## Data Availability

All data produced in the present study are available upon reasonable request to the authors.

https://atheroexpress.nl/data-collection/

